# Deep skin dysbiosis in vitiligo patients: link with mitochondrial and immune changes

**DOI:** 10.1101/2020.07.29.20163469

**Authors:** Hanene Bzioueche, Kotryna Simonyté Sjödin, Christina E West, Abdallah Khemis, Stéphane Rocchi, Thierry Passeron, Meri K Tulic

**Affiliations:** Université Côte d’Azur, INSERM U1065, Centre Méditerranéen de Médecine Moléculaire (C3M), Nice, France; Department of Clinical Sciences, Pediatrics, Umeå University, Umeå, Sweden; Côte d’Azur University. Department of Dermatology, University Hospital of Nice, France

**Author notes:** corresponding authors **Correspondences:** Dr Meri K Tulic, INSERM U1065, C3M, 150 route de Ginestiere 06200 Nice, France, Pr. Thierry Passeron, Department of Dermatology, Archet 2 Hospital., 150 route de Ginestiere 06200 Nice, France. These two authors contributed equally to this work (Hanene Bzioueche, Kotryna Simonyté Sjödin). equal senior-author contributions (Thierry Passeron, Meri K Tulic).

**Keywords:** skin dysbiosis, microbiota, vitiligo, immunity, melanocytes

## Abstract

**Rationale:** Vitiligo is an autoimmune-disease characterized by patchy, white skin due to melanocyte loss. Commensal cutaneous or gut dysbiosis have been linked to various dermatological disorders. Here, we studied skin and gut microbiota of vitiligo patients compared to healthy controls.

**Methods:** We recruited 20 subjects and obtained swabs and biopsies from lesional and non-lesional skin, stool and blood from each individual (total 100 samples).

**Results:** We detected reduced richness and distribution of microbiota in stool of vitiligo subjects compared to controls (P<0.01). Skin swabs had greater alpha-diversity than skin biopsies (P<0.001), however only trends were seen between groups when examining microbiota at the skin surface. This was in contrast to sampling deeper layers of skin from the same patients which showed decreased richness and distribution of species (P<0.01) but greater phylogenetic diversity (P<0.01) in lesional compared to non-lesional sites. Biopsy microbiota from the lesional skin had distinct microbiota composition which was depleted of protective *Bifidobacterium* and enriched in *Terenicutes, Streptococcus, Mycoplasma* and mitochondrial DNA (P<0.001); the latter was linked with increased innate immunity and stress markers in the blood of the same patients (P<0.05).

**Conclusion:** These data describe vitiligo-specific cutaneous and gut microbiota and, for the first time in humans, a link between mitochondrial alteration, innate immunity and skin microbiota.

## INTRODUCTION

Vitiligo is characterized by an acquired loss of melanocytes in the skin and sometimes in the hair follicles. It is now well demonstrated that CD8+ T cells attracted in the epidermis are responsible for the melanocytic loss. Genome-wide association studies (GWAS) identified over 50 vitiligo susceptibility loci involved in melanogenesis and immunity in vitiligo patients [1]. However, these genetic studies and a delay in vitiligo age-of-onset over the past 30 years emphasize the key role of environmental factors in triggering vitiligo [2]. We recently demonstrated that vitiligo patients have an increased number of natural killer (NK) and innate immune cells (ILC1) in their non-lesional skin and their blood, and that those innate cells have a greater response to stress, producing higher amount of IFNγ leading to the production of chemokine (C-X-C motif) ligand 9 (CXCL9), CXCL10 and CXCL11 by both keratinocytes and melanocytes and to a CXCR3B-induced apoptosis in melanocytes [3]. We demonstrated that these early events which lead to CD8+ T cell-mediated adaptative response against melanocytes can be triggered by endogeneous or exogeneous stress, namely damage and pattern-associated molecular patterns (DAMPs and PAMPs, respectively). Bacteria are among the top producers of PAMPs and could participate in activation of the innate immune response in vitiligo. Interestingly, gut dysbiosis has been reported in several auto-immune disorders. Although less reported, skin dysbiosis is also observed in some dermatoses, such as acne, psoriasis and atopic dermatitis [4–6]. Most of these studies assessed the superficial microbiome using swabs. However, recent data emphasized the differences in the microbiome between the surface and the deeper part of the skin using biopsy samples [7,8]. Surprisingly, data on the microbiome in vitiligo patients remains sparse. To date, there exists only one study suggesting dysbiosis in vitiligo skin [9], however the authors only assessed the superficial skin microbiota using swabs and compared lesional to non-lesional vitiligo skin without including healthy controls. A more recent study in a mouse model of vitiligo has shown that antibiotic-induced depletion of *Bacteroides*-dominated population in the gut may induce depigmentation in the skin suggesting a possible link between gut and skin compartments [10]. In the present study, we compared for the first time, the microbiota of gut, superficial and deeper parts of lesional and non-lesional skin of vitiligo patients and compared these results to sample and site matched healthy controls. We found gut dysbiosis with a reduced *Bacteroides* population in vitiligo patients. We also report marked differences of the microbiota using swabs and skin biopsies. Finally, we report significant variation in microbiota diversity and quality in the deeper regions of lesional vitiligo skin and these changes to be associated with mitochondrial damage and changes in skin immunity.

## RESULTS

### Vitiligo disease is associated with lower α-diversity and increased Firmicutes/Bacteroidetes fraction in the gut

First, we set out to examine if we could detect differences in microbial composition and diversity in the gut of patients with vitiligo skin disease (n=10) compared to healthy controls (n=10). Here we show lower gut microbial α-diversity in vitiligo patiens compared to healthy controls (for observed OTUs P=0.005; Shannon diversity index P=0.115; and Faith’s Phylogenetic Diversity H=5.851, P=0.006) (Figure 1A). β-diversity did not differ between the groups. We fousnd trends for higher bacterial relative abundance (RA) of *Firmicutes* (P=0.09) and lower *Bacteroidetes* (P=0.08) in the stools of vitiligo subjects compared to healthy controls (Figure 1B and Supplementary Table 1) which is likely to be a result of reduced *Bacteroides* (Figure 1C and Supplementary Table 2). The ratio between *Firmicutes* and *Bacteroidetes* (F/B) was 8.5±1.2 (mean±SEM) controls and 18.5±2.6 for vitiligo patients (P<0.01).

**Figure 1.**
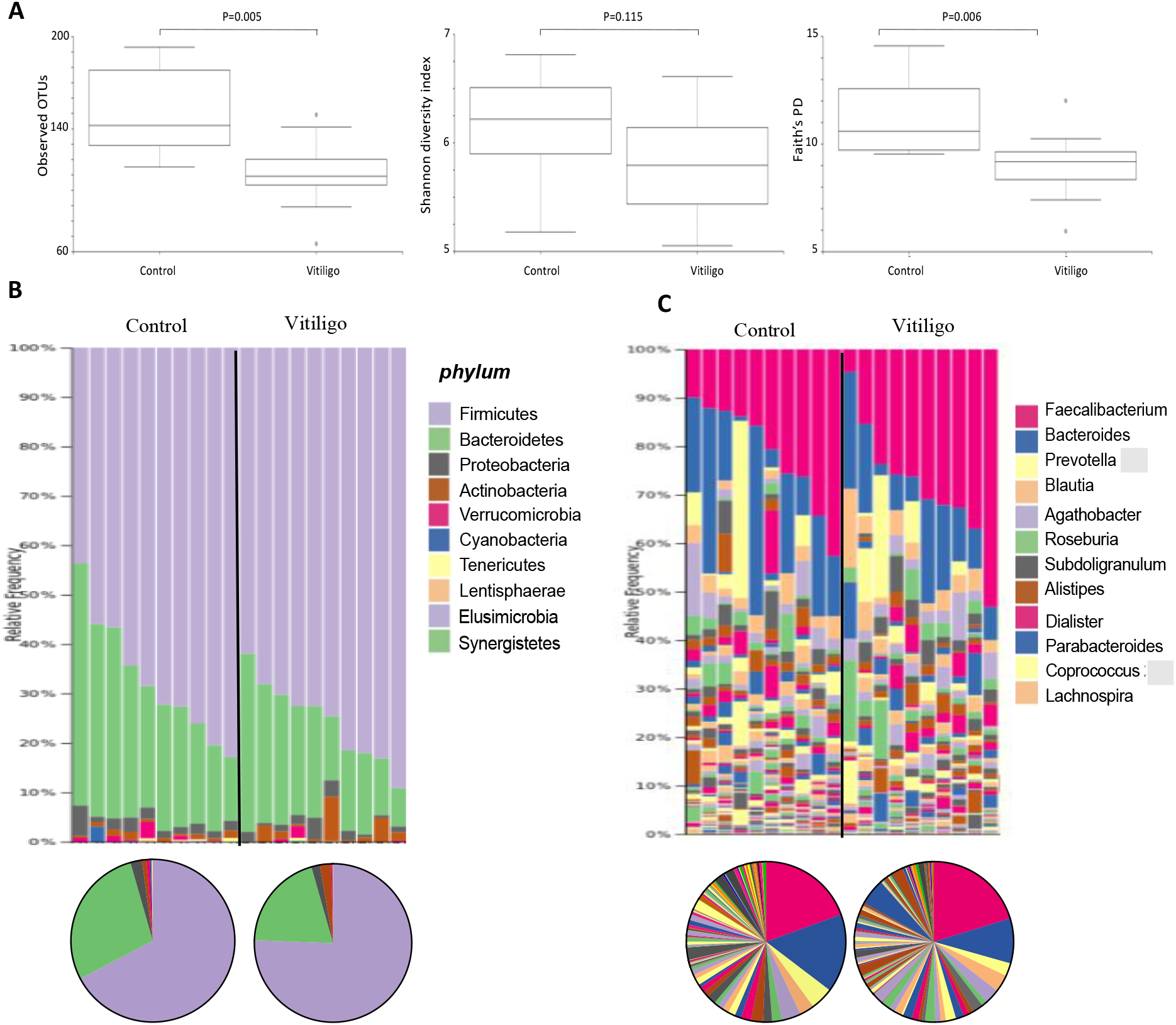
Reduced α-diversity and increased Firmicutes/Bacteroides ratio in the gut microbiota of vitiligo subjects. **(A)** Boxplots illustrating the comparison of different measures of **α**-diversity between healthy (n=10) and vitiligo (n=10) stool samples. **(B)** Bacterial composition and diversity expressed as a percentage of relative abundance between the two groups at the phylum and **(C)** genus levels. Individual subjects are shown as taxa bars (above) and grouped data as pie charts (below). A complete list of OTUs is shown in Supplementary Table 1 and Table 2.

### Significant microbial dissimilarity between swabs and biopsies

To examine if there are differences in microbial composition and diversity in the skin of vitiligo patients compared to healthy skin, we studied microbiota extracted from paired skin swabs and skin biopsies from the same subject as well as collected samples from the patient’s lesional and non-lesional skin sites. We compared microbial communities α- and β-diversity between swab or biopsy samples from control, lesional (L) and non-lesional (NL) sites (n=30 swab and n=30 biopsy). Observed OTUs and Shannon diversity index were higher in swabs compared to biopsies (P<0.001 for both; Figure 2A), whereas Faith’s PD was higher in biopsies (P<0.001; Figure 2A), the latter suggesting higher phylogenetic richness in biopsies compared to swabs. These results suggest richness and distribution of bacteria (α-diversity) differs in relation to the sampling method. Furthermore, our results demonstrate distinct and significant differences in ß-diversity between biopsies and swabs (P<0.001, Figure 2B). We found higher *Bifidobacterium* (with *B. bifidum* and *B. longum*), *Escherichia-Shigella, Parabacteroides* and *Enterococcus* in biopsies compared to swabs, whereas *Staphylococcus, Paracoccus, Kocuria, Micrococcus* and *Anaerococcus* were more abundant in the skin swabs (p<0.05 for all comparisons; Figure 2C). In the swabs, *Staphylococcus* was the most abundant genus making up > 30% of all bacteria while in the biopsies, *Bifidobacterium* was the most common, making up almost 60% of the total bacterial load. Taken together, these results demonstrate a marked difference in the skin microbiome when analyzed using swab *versus* biopsy sampling and there are several bacterial genera that correlates strongly and significantly with the sample collection method or the depth of microbiota sampling.

**Figure 2.**
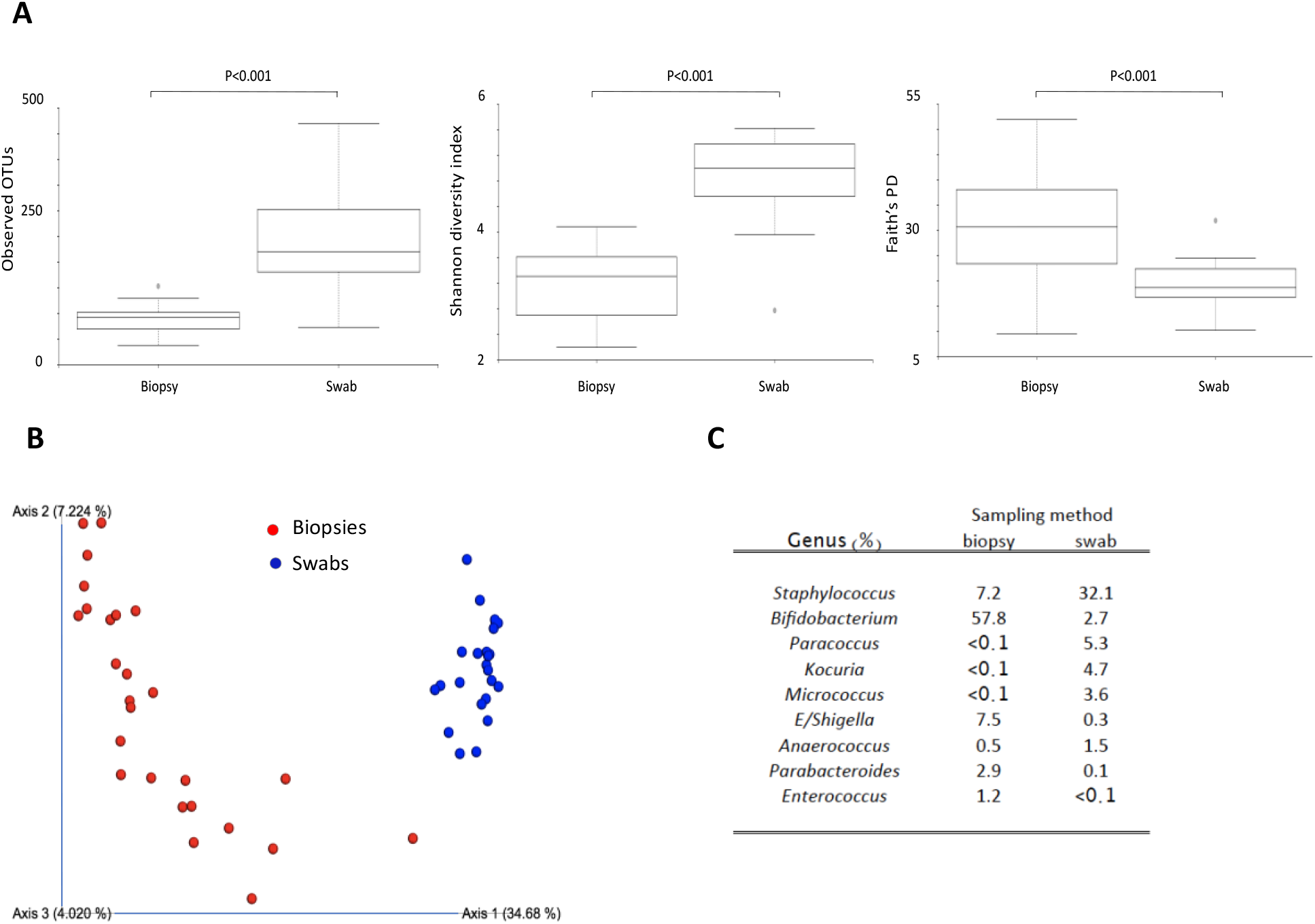
Difference in bacterial diversity and bacterial composition between swabs and biopsies due to sampling method. (**A)** Boxplots illustrating the comparison of different measures of α-diversity in paired samples of skin biopsies (n=30) and skin swabs (n=30). **(B)** PCoA plot representing β-diversity in biopsies (red circles) and swabs (blue circles) (p<0.001). (**C**) Demonstrates sampling-method related differential bacterial abundance where bacteria enriched at each site is denoted and given as a percentage of total genus.

### Depletion of Staphylococcus and enrichment of Proteobacteria in lesional swabs of vitiligo patients

Swab samples from NL and L vitiligo skin had a significantly higher α-diversity compared to healthy control swabs (P<0.05) (Figure 3A). There was no difference in α-diversity between NL and L skin samples (Figure 3A). We found no significant differences in β-diversity between healthy, NL and L skin swab samples. We observed decreased *Firmicutes* in L compared to NL sites (p<0.01) (Figure 3B and Supplementary Table 3); most likely due to decreased *Staphylococcus* at this site (p<0.02) (Figure 3C and Supplementary Table 4). Examining the bacterial composition of swab microbiota at the genus level, we show that greater than half of taxonomic assignments on skin surface belong to 5 major genera: *Staphylococcus, Corynebacterium, Cutibacterium* (formerly *Propionibacterium*), *Kocuria* and *Paracoccus* (Figure 3C). We found a trend for lower *Cutibacterium* in L compared to NL swabs (p=0.057) (Figure 3C) which may have important implications in vitiligo as the loss of *Cutibacterium* diversity triggers activation of the innate immune system [11]. We have also seen trends for increased *Proteobacteria* (p=0.08) phylum in L compared to NL swabs (Figure 3B). As *Proteobacteria* contains many well-known human pathogens and is related with disbalance and inflammation [12], we investigated this phylum in more detail. We have shown a disease-dependent increase in presence of *Gammaproteobacteria*, that is relative abundance in L>NL>C swabs and disease-dependent decreased in *Alphaproteobacteria* (L<NL<C) (P=0.046) (Supplementary Figure 1A and Supplementary Table 5). Lesional skin enrichment with *Gammaproteobacteria* has also been shown in the only other study to date in vitiligo subjects [9]. We have shown trends for increased *Paracoccus* (p=0.09) and *Acinetobacter* (p=0.08) in vitiligo swabs compared to control swabs (Supplementary Figure 1B). Differences in *Haematobacter* were not statistically different due to patient variability.

**Figure 3.**
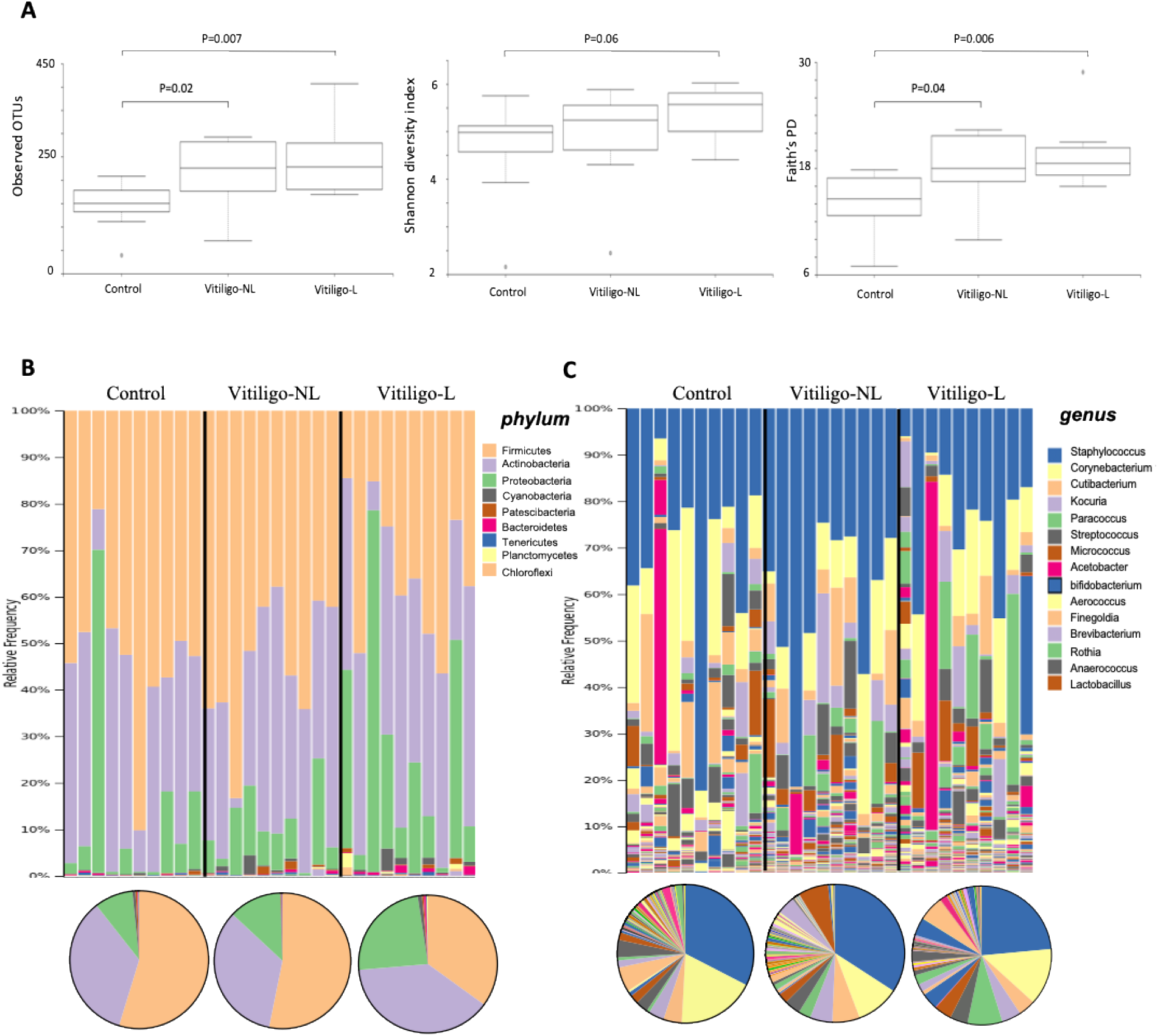
Higher α-diversity, depletion of Staphylococcus and enrichment of Proteobacteria in lesional swabs of vitiligo patients. (**A**) Boxplots illustrating the comparison of different measures of α-diversity in skin swabs taken from non-lesional (NL) and lesional (L) regions of the skin from vitiligo patients (n=10) compared to skin swabs taken from healthy controls (n=10). (**B**) Bacterial composition and diversity (expressed as percentage of relative abundance) between the groups are shown at the phylum and (**C**) genus levels with individual taxa data shown above and grouped average data as pie charts below. A complete list of OTUs is shown in Supplementary Table 3 and Table 4.

### Biopsy microbiota in lesional skin have distinct composition and are associated with mitochondrial damage

In contrast to the swabs, microbial community β-diversity in L skin biopsies differed significantly compared to NL skin biopsies or healthy controls (p<0.001) (Figure 4A) and had the most distinct microbiota compared to all other groups (Supplementary Figure 2). We saw a decrease in species richness and distribution (Shannon index, P=0.006) but increase in phylogenetic diversity (Faith’s PD, P=0.003) in biopsies from L compared to NL skin, whereas Shannon index was higher in NL skin compared to controls (P<0.01) (Figure 4B). The taxa bar plots suggest decreased *Actinobacteria* (p=0.09) and increased *Firmicutes* (p=0.08) in L compared to NL or control biopsies (Figure 4C and Supplementary Table 6). In our cohort, the largest difference was the increase in *Tenericutes* (P<0.001 for control versus NL and P<0.02 for NL versus L) and mitochondrial DNA (p<0.007) exclusively in biopsies from 7/10 lesional skin samples whereby 3 out of the 10 samples looked like NL skin (Figure 4C).

**Figure 4.**
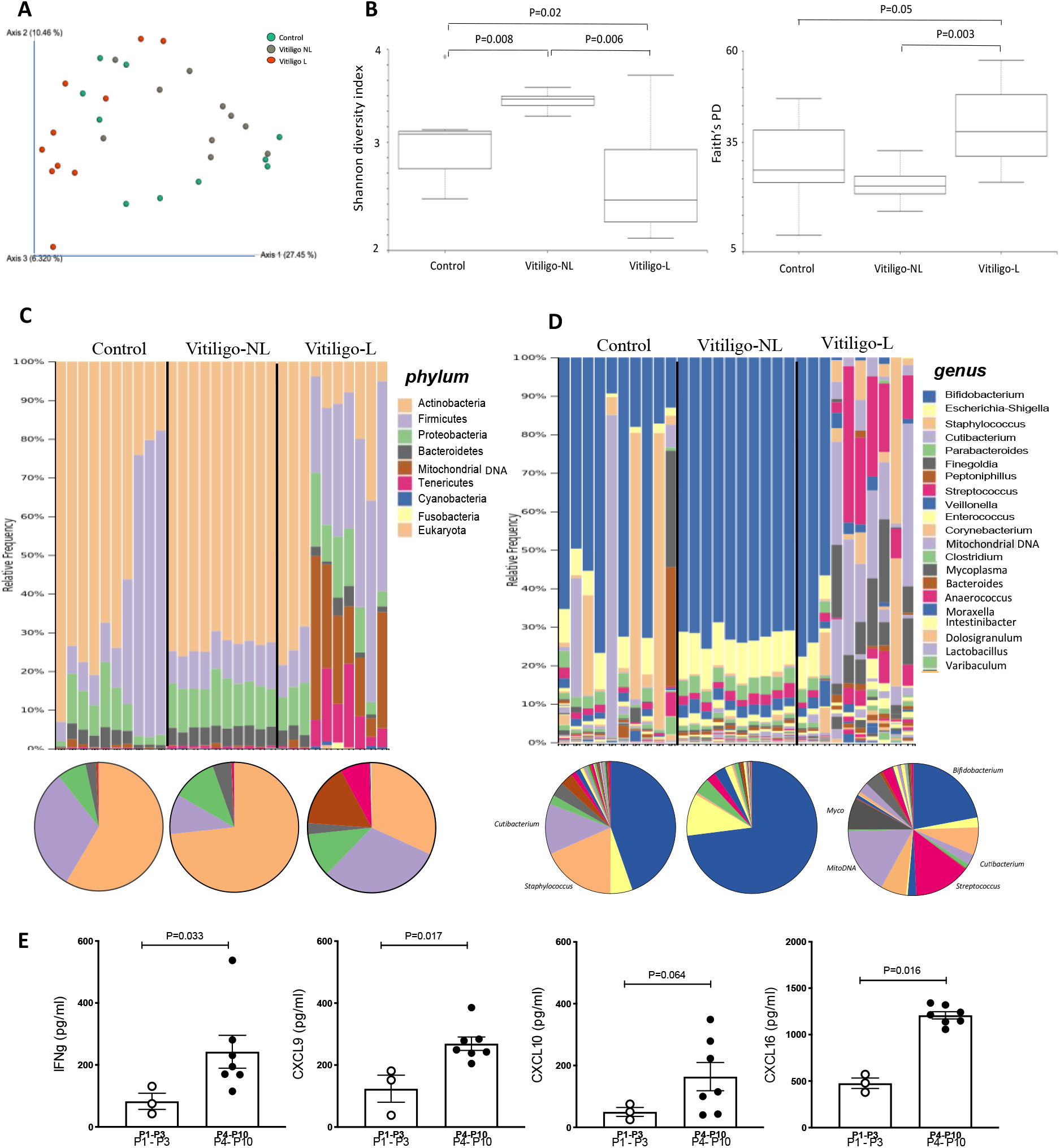
Lesional biopsies from vitiligo patients have distinct microbiota and are associated with changes in bacterial diversity, mitochondrial damage and activation of the innate immune system. **(A)** PCoA plot representing beta diversity between the different groups illustrating biopsies from the lesional zone of vitiligo patients (red circles) to be significantly different from all other samples (P<0.001). (**B**) Boxplots illustrating the comparison of different measures of α-diversity in skin biopsies taken from non-lesional (NL) and lesional (L) regions of the skin from vitiligo patients (n=10) compared to skin biopsies taken from healthy controls (n=10). (**C**) Bacterial composition and diversity (expressed as percentage of relative abundance) between the two groups at the phylum and genus **(D)** level. Individual subjects are shown as taxa bar plots (above) and grouped data as pie charts (below). A complete list of OTUs is shown in Supplementary Table 6 and Table 7. **(E)** Measurements of innate (IFNγ, CXCL9, CXCL10) and stress marker (CXCL16) proteins in the serum of vitiligo patients with elevated mitochondrial signature (patients 4-10 in C) compared to vitiligo patients without increased mitochondrial signature (patients 1-3).

In the skin biopsies, greater than half of taxonomic assignments belong to 5 major *genera* (3 overlapping with the swabs): *Bifidobacterium, Escherichia-Shigella, Staphylococcus, Cutibacterium* and *Parabacteroides* (Figure 4D and Supplementary Table 7). The heat map illustrates the top 20 most differentially expressed bacterial taxa between the three groups at the individual level (Supplementary Figure 3). Pair-wise analysis for differential abundance between NL vitiligo biopsies and control biopsies have shown *Staphylococcus* and *Cutibacterium* to be depleted in NL biopsies (p=0.07); the same two *genera* that were depleted on the surface of L skin. In contrast, *Enterococcus, Mycoplasma, Veillonella, Intestinibacter, Bacteroides, Escherichia-Shigella, Parabacteroides, Bifidobacterium*, and *Streptococcus* were enriched at NL sites (all p<0.007) (Supplementary Figure 3). Comparisons between L and NL biopsies have shown L skin to be further enriched for *Streptococcus* as well as *Proteobacteria* including *Gemella, Raistonia* and *Undibacterium* (all p<0.05) with a large and specific increase in *Mycoplasma* and Mitochondrial DNA (Figure 4D). In contrast, *Bifidobacterium, Escherichia-Shigella, Parabacteroides* and *Enterococcus* were significantly depleted taxa in biopsies from L compared to NL skin and compared to healthy biopsies (all p<0.05). *Bifidobacterium* was completely depleted in 7/10 L biopsies when compared to their matched NL sites in which *Bifidobacterium* makes up almost 60% of the total bacterial genus (Figure 4D).

Finally, we further investigated the differences between the first 3 patients who didn’t have an increase in mitochondrial DNA in their skin biopsy to the remaining 7 patients who did. Interestingly, these 3 patients had a similar microbiome to non-lesional samples, that is, low abundance of *Tenericutes* and *Firmicutes* with predominant presence of *Actinobacteria* (Figure 4C). The difference between these 2 sub-groups of patients could not be explained by sex, age, location of the biopsies, disease activity or disease duration. To further investigate if these changes in the skin could activate the innate immune response, we measured the levels of IFNγ and chemokines CXCL9 and CXCL10 that we previously have shown to be increased after stimulation of the innate cells of vitiligo patients by DAMPs or PAMPs [3]. We also measured the levels of CXCL16 that is produced by keratinocytes and mediates CD8+ T cell skin trafficking under oxidative stress in vitiligo patients [13]. The 7 patients with increased mitochondrial DNA in lesional sites had a significantly increased levels of IFNγ, CXCL9 and CXCL16 (P<0.05) in their serum compared to the patients without elevated mitochondrial DNA (Figure 4E). The corresponding levels of CXCL-10 in the serum were of borderline significance (P=0.064).

## DISCUSSION

Gut, and to a lesser extent, skin dysbioses have been reported in many inflammatory and autoimmune disorders. Surprisingly, microbiome studies in vitiligo remain limited. Here we have shown, for the first time, dysbiosis in the gut and skin microbiota of vitiligo subjects compared to controls. The most striking differences were detected in the deep dermis layer of the lesional skin where enrichment of pathogenic *Tenericutes* bacteria and a loss of protective *Bifidobacterium* were associated with mitochondrial damage at the same site as well as increased stress and immune activation markers in their peripheral blood. We do acknowledge the relative limited number of subjects analysed being a potential weakness of this study. It is important to state that this was an invasive study requiring six samples *per* patient (2 skin biopsies, 2 skin swabs, stool and a blood sample). Moreover, the group numbers were based on recent human microbiome studies using the similar sequencing approach [9,14–17].

We detected reduced *α*-diversity (richness and distribution of bacterial species) in the stool of vitiligo compared to controls with increased abundance of *Firmicutes* and reduced *Bacteroidetes*. The ratio between *Firmicutes* and *Bacteroidetes* (F/B) was significantly higher for vitiligo patients. A similar increase in the F/B ratio has previously been reported in other autoimmune diseases such as type I diabetes, Grave’s disease, lupus and multiple sclerosis as well as autoimmune skin conditions including scleroderma and psoriasis [18]. Interestingly, a protective effect of *Bacteroides* was recently reported in the gut of a mouse vitiligo model and was associated with skin depigmentation of mice [10]. If the same protection holds true for human vitiligo, reduction and/or lack of protective *Bacteroides* in the human gut may contribute to disease onset or exacerbation. It is important to note that vitiligo depigmentation is associated with better prognosis in melanoma patients [19] and interestingly, our stool microbiota results in vitiligo patients (high F/B ratio) have striking similarities with good responders in melanoma patients treated with anti-PD1 [20]. Together, these results demonstrate differences in gut microbial composition between vitiligo and healthy controls and may suggest a possible gut-skin axis in this disease.

Our results analyzing skin microbiota emphasize a marked difference in the skin microbiota when analyzed using swab *versus* biopsy sampling suggesting that there are several bacterial genera that correlates strongly and significantly with the sample collection method. Most of the studies to date that assessed the microbiota in skin disorders have used only swab samples. The unique aspect of our study is that we obtained stool, skin swab and skin biopsy samples from the same individuals thereby reducing inter-patient variability and strengthening robustness of our study. Our results are in agreement with those of Prast-Nielsen [7] and emphasize the need for simultaneous collection of swab (surface) and biopsy (deep skin) samples as biopsies are providing additional information which is critical in correct interpretation of the data. Using swab samples, we found some albeit small differences and trends in differential abundance of microbial composition between healthy and vitiligo skin swabs; the largest differences seen in depletion of commensal *Staphylococcus* and *Acinetobacter* and enrichment of pathogenic *Paracoccus* (*Proteobacteria)* in lesional skin. *Staphylococcus epidermidis* is a known important symbiont in the skin which directly interacts with cutaneous immune cells to regulate skin homeostasis [21]. *Staphylococcus* spp are also known to colonise lesions in a number of dermatological diseases including atopic dermatitis and presents propensity to form biofilms; adhesive surface-attached colonies that become highly resistant to antibiotics and immune responses [22].

Current understanding of the skin microbiota is based on swab sampling the outermost layer of the epidermis. Despite the invasive nature of biopsies, sampling dermal microbiota has become critically important as the dermal community is less affected by external factors [8]. Our results give us, for the first time, a comprehensive profiling of the microbial species that are depleted in full thickness subepidermal compartments of the vitiligo skin which may be targeted for therapy. Thus, in contrast to the swabs, marked differences were found in microbial community in lesional skin biopsies compared to non-lesional skin biopsies or healthy controls with a decreased *Actinobacteria* and increased *Firmicutes*. Interestingly, the same imbalance between *Actinobacteria* and *Firmicutes* in lesional *versus* non-lesional skin was found in the only other study that assessed skin microbiota in vitiligo skin [9]. However, in our cohort, the largest difference was the increase in *Tenericutes* and mitochondrial DNA in biopsies from lesional skin. With such ‘black-and-white’ results even with the limited number of subjects, it is tempting to speculate that *Tenericutes* may be used as a marker phylum for patients that may benefit from a supplementation of probiotics or bacterial lysates with bifidobacteria in addition to established treatment. Further analyses and interventional studies with bifidobacteria supplementation are required to confirm this hypothesis.

The latest evidence in the literature emphasizes the interaction between mitochondria and microbiota [23] ^19^. It has been recently shown that the mitochondrial genotype modulates both reactive oxygen species (ROS) production and the species diversity of the gut microbiome [24]. Interestingly, mitochondrial alterations with increased production of ROS have been reported in vitiligo cells [25,26]. Our results are in accordance with these *in vitro* studies and demonstrate potent mitochondrial damage in lesional skin of patients. As observed in the gut of mice [24], α-diversity decreased in samples with high mitochondrial DNA. Modification of the microbiome only in the biopsy samples having mitochondrial DNA strongly argues in favor of microbiome modification induced by the mitochondrial stress. Interestingly, the genus that was almost completely absent in the 7 patients with the mitochondrial DNA in the skin is *Bifidobacterium* which is well-known to have protective effects and to decrease the activation of innate immune cells [27]. Moreover, those 7 patients had also a significantly increased levels of IFNγ, CXCL9, CXCL10 and CXCL16 in their serum compared to the patients without elevated mitochondrial DNA. Thus, we can hypothesize that similarly to what has been shown in the gut of mice [24], primitive mitochondrial stress in some vitiligo patients could be responsible for the change in the skin microbiome. This skin dysbiosis could then trigger the innate response through PAMPs and lead to the production of IFNγ and chemokines that can be detected in the serum of these patients. Of course, this hypothesis needs to be confirmed in larger number of patients and ideally metagenomics should be performed to characterize the origin of mitochondrial DNA.

There are a number of unique aspects to this study. First and foremost it is its novelty. To our knowledge, there are no data examining the role of microbiota in vitiligo skin compared to healthy skin. Secondly, there are matched multiple samplings (swab, biopsy, stool, serum) from each patient avoiding inter-patient variability and thereby allowing for robustness of the data even with a relatively limited sample size. Our key data demonstrating differences between skin swabs and skin biopsy microbiota and between lesional compared to non-lesional or control skin are all highly significant (P<0.001) suggesting important biological differences between these sites. Thirdly, we have demonstrated the skin dysbiosis in vitiligo patients is associated with profound mitochondrial damage and loss of protective bacteria at the same site with heightened innate immune response in the blood of vitiligo patients compared to healthy controls. These novel data not only describes vitiligo-specific cutaneous and gut microbiota, but it also shows, for the first time in humans, a link between mitochondrial alteration, microbiota and innate immunity. This opens up new avenues to explore in potential treatment of vitiligo and other auto-immune disorders.

## MATERIALS AND METHODS

### Biological samples

Twenty (20) subjects were recruited for this study (Table 1). Ten vitiligo patients with stable disease and 10 healthy patients were recruited from the Department of Dermatology, Archet 2 Hospital, CHU Nice, after informed, written consent was obtained. Exclusion criteria included presence of other autoimmune diseases or *prior* treatment (phototherapy, topical or systemic corticosteroids, systemic antibiotics and/or other immunosuppressive agents) for up to 3 months before skin sampling. The groups were matched for gender, age, and biopsy location. All skin samples were taken from the dry locations (Table 1). Swab and biopsy samples were collected at the same time for each patient. From each vitiligo patient, we obtained two 4-mm skin punch biopsies (1 from lesional, L and 1 from non-lesional, NL site) and from each healthy patient 1 skin biopsy of the same size. Skin swabs were taken at the same time from adjacent skin area sampling 5cm × 5cm L and NL skin sites using sterile dry swabs (Copan Diagnostics) and rinsed in 200 µl sterile 0.15 M NaCl and 0.1% Tween20 (v/v). The same morning, stool samples as well as 10 mL of blood was also collected from all subjects. Serum was obtained from the blood following Ficoll gradient centrifugation (Lymphoprep®, Euromedex, France). All biological samples (n=60 from vitiligo patients and n=40 from healthy controls) were stored at -80° C until analysis. The study was approved by the National Ethics Committee (N14.028) and was carried out in accordance with The Code of Ethics of the World Medical Association (Declaration of Helsinki).

**TABLE 1.** Patient Characteristics.

| **Characteristics** | **Healthy controls** | **Vitiligo** |
| --- | --- | --- |
| *Subjects (n)* | 10 | 10 |
| *Age (years), mean (SE)* | 60.2 (2.8) | 51.5 (3.9) |
| *Gender (male/female)* | 4/6 | 3/7 |
| *Mean disease duration (years)* | - | 3.2 (1.4 - 22) |
| *Family history (%)* | - | 2 (20%) |
| *Location of biopsy (n)* | Shoulder (5), forearm (3), leg (1), ankle (1) | Forearm (5), leg (4), ankle (1) |

### DNA isolation from stool samples

Eighty to 120 mg of frozen stool was transferred to Precellys soil grinding SK38 lysing tubes (Bertin Technologies, Montigny-le-Bretonneux, France) and one volume of warm (37° C) lysis buffer [4% (w/v) SDS, 50 mM TrisHCl pH 8.0, 500 mM NaCl, 50 mM EDTA] was added. Samples were homogenized for 10 mins at room temperature using a Vortex adapter. Lysozyme (Sigma Aldrich Chemie GmbH, Germany; final concentration 6.25 mg/ml) was added and samples were incubated at 37° C for 30 min, then transferred to 80° C heating block and incubated for 15 mins by inverting every 5 mins. Samples were then centrifuged at 4° C 20,000g for 5 mins and supernatants were collected; proteinase K (Roche Diagnostics GmbH, Germany) was added (final concentration 0.4 mg/ml) and the samples were incubated on a heating block at 70° C for 10 mins. After incubation, 10 M NH_4_OAc (final concentration 2 M) was added and samples were incubated on ice for 5 mins, then centrifuged at 4° C 20,000g for 10 mins. Supernatants were collected and an equal volume of cold isopropanol was added. The samples were stored on ice for 30 mins and thereafter centrifuged at 4° C 20,000g for 20 mins. Pellets were washed 2-3 times with cold 70% ethanol, dried and dissolved overnight in commercial Tris-EDTA buffer (1xTE). The following day, the DNA concentrations were measured using Qubit dsDNA Broad Range Assay kit (Thermo Fisher Scientific Inc., USA) on Qubit 3.0 fluorometer (Thermo Fisher Scientific Inc., USA). RNAse (Thermo Scientific, Lithuania) at final concentration of 1 µg/µl.

### DNA isolation from skin samples

Bacterial DNA was extracted from skin biopsies and swabs using QIAamp□ DNA Microbiome Kit (Cat. No. 51704) (QIAGEN GmbH, 40724 Hilden, Germany). We followed manufacturer’s protocol with a single modification. After addition of Proteinase K the second time, samples were incubated overnight on a heating block at 56° C. After eluting DNA, the concentrations were measured using QubitTM 1X dsDNA HS Assay Kit (Life Technologies Corporation, Oregon 97402, USA) on Qubit 4.0 fluorometer (Thermo Fisher Scientific Inc., USA).

### 16S rRNA Gene Library Preparation and Amplicon Sequencing

The sequencing library was prepared according to Earth Microbiome Project’s Protocol [28] with the following modifications. The fused primers were modified to contain barcode sequence on both forward (341F) and reverse (805R) primers and selected to target the V3-V4 region instead of the V4 region. Sequences of fused primers are provided as Supplementary Table 8. The PCR reactions for library preparation were carried out in triplicates as follows: 20 ng of template DNA was mixed with 5PRIME HotMasterMix (Quantabio, USA) consisting of: 1U Taq polymerase, 45 nM Cl, 2.5 mM Mg2+, 0.2 mM of each dNTP, 0.2 µM of each primer (Eurofins Genomics, Germany) and 0.64 ng bovine serum albumin (BSA) in a final volume of 25 µl *per* reaction. The PCR conditions were: 90° C for 15s and 94° C for 3 mins followed by 35 cycles of 94° C for 45s, 50° C for 1 min and 72° C for 1.5 mins, after which a final elongation step at 72° C for 10 mins was performed. The triplicates were pooled and visualized on 1% agarose gel to estimate the size of the amplicons. DNA concentrations of the amplicons were measured as described in the section above. Every PCR run includeed a negative (water) and a positive (MOCK community sample) control. The following reagent was obtained through the NIH Biodefense and Emerging Infections Research Resources Repository, NIAID, NIH as part of the Human Microbiome Project: Genomic DNA from Microbial Mock Community B (Even, Low Concentration), v5.1L, for 16S RNA Gene Sequencing, HM-782D. Libraries were then pooled in equimolar concentrations and the amplicon pool was purified according to protocol using AMPure XP beads (Beckman Coulter, USA). Prior to amplicon sequencing the amplicon pool was diluted in 10 mM Tris-HCl (pH 8.5) to a final concentration of 5 nM. Following the Illumina recommendations, the amplicon pool was denaturated using an equal amount of 0.2 M NaOH (BioUltra) (Sigma Aldrich Chemie GmbH, Germany) and further diluted to 12 pM in hybridization buffer (HT1 buffer included in the Reagents Kit v3, Illumina, USA). The pool was finally spiked with 5% denaturated PhiX control library (Illumina, USA). The sequencing was performed using the MiSeq sequencing platform with the Reagents Kit v3, 600 cycles (Illumina, USA).

### Microbiome Analyses: Composition, Diversity and Discovery of Metagenomic Biomarkers

The composition and diversity of the gut microbiome were assessed using QIIME2 [29]. Initially, read-pairs were demultiplexed using deML [30] before sequences were quality filtered and denoised using DADA2 [31]. For stool samples, total sequencing yield was 1,595,147 reads with median of 74,594 (n=21). For skin samples 7,499,237 reads was a total yield distributed among 73 samples, with median of 96,497. Raw sequence data were demultiplexed and quality filtered using the q2-demux plugin followed by denoising with DADA2 [31]. All amplicon sequence variants (ASVs) were aligned with mafft [32] and used to construct a phylogeny with fasttree2 [33]. *α*-diversity metrics (observed OTUs, Shannon index and Faith’s Phylogenetic Diversity [34]) and β-diversity metrics (unweighted UniFrac) [35] and also Principle Coordinate Analysis [PCoA]) were estimated after samples were rarefied to 1000 sequences *per* sample. Shannon index accounts for both richness (number of species) and distribution (how evenly the species are distributed) while Faith’s PD pertains more to phylogenetic diversity in a sample. β-diversity measures differences in microbial composition between different samples. Taxonomy was assigned to ASVs using the q2-feature-classifier [36] classify-sklearn naïve Bayes taxonomy classifier against the SILVA full-length 99% OTUs reference sequences [37].

### ELISA

IFNγ, CXCL9 and CXCL10 were measured from the serum using commercially available ELISA kits (Peprotech, NJ, USA) while CXCL16 ELISA was purchased from R&D Systems (Minnesota, USA).

### Statistical analyses

Differences in α- and β-diversity between groups were calculated using Kruskal-Wallis test (p value is FDR adjusted) and PERMANOVA (number of permutations=999), respectively. We have used ANCOM [38] to investigate differentially abundant bacteria between the groups (P<0.05, FDR adjusted for the analysis). Additionally, non-parametric independent sample t-test (between controls and vitiligo subjects) and paired sample t-test (between lesional and non-lesional samples) were used. For all tests P<0.05 was considered significant and P between 0.06 and 0.1 a trend.

## Data Availability

Sequence data have been deposited to the Sequence Read Archive (SRA) under accession number PRJNA639425. All other data that support the findings of this study are available from the corresponding authors upon reasonable request.

## ACKNOWLEDGEMENTS

We would like to thank Mona Svensson and Carina Lagerqvist for laboratory work, Emelie Näslund Salomonsson for assistance with Illumina MiSeq run, as well as Sonia Amroune and Raja Bahroumi for their assistance in clinic with patient recruitment. This work was supported by the Institut National de la Santé et de la Recherche Médicale (INSERM).

## AUTHOR CONTRIBUTIONS

HB was involved in sample collection and their processing, data analysis and interpretation, as well as manuscript writing and construction of Figures. KSS along with CEW optimized the protocol for extraction of DNA from skin biopsies. KSS performed DNA extractions, 16S sequencing, statistical analysis and construction of Figures. KSS, SR and CEW provided intellectual input, expertise and feedback on manuscript draft. AK performed all swab and biopsy collections from vitiligo patients. TP was involved in patient recruitment. MKT performed all the ELISA measurements. MKT and TP conceived the experiments, obtained financial support, supervised the study, were involved in data interpretation and together wrote the manuscript. All authors have read and approved the final version of the paper.

## REFERENCES

1. Roberts GHL, Santorico SA, Spritz RA. The genetic architecture of vitiligo. Pigment Cell Melanoma Res. 2020; 33:8–15.

2. Jin Y, Santorico SA, Spritz RA. Pediatric to Adult Shift in Vitiligo Onset Suggests Altered Environmental Triggering. J Invest Dermatol. 2020; 140:241–243.

3. Tulic MK, Cevazza E, Cheli Y, Jacquel A, Luci C, Cardot-Leccia N et al., Innate lymphocyte-induced CXCR3B-mediated melanocyte apoptosis is a potential initiator of T-cell autoreactivity in vitiligo. Nature Comm. 2019; 10:2178.

4. O’Neill AM, Gallo RL. Host-microbiome interactions and recent progress into understanding the biology of acne vulgaris. Microbiome. 2018; 6:177.

5. Stehlikova Z, Kostovcik M, Kostovcikova K, Kverka M, Juzlova K, Rob F et al. Dysbiosis of Skin Microbiota in Psoriatic Patients: Co-occurrence of Fungal and Bacterial Communities. Front Microbiol. 2019; 10:438.

6. Williams MR, Gallo RL. Evidence that Human Skin Microbiome Dysbiosis Promotes Atopic Dermatitis. J Invest Dermatol. 2017; 137:2460–2461.

7. Prast-Nielsen S, Tobin AM, Adamzik K, Powels A, Hugerth LW, Sweeney C et al. Investigation of the skin microbiome: swabs vs. biopsies. B J Dermatol. 2019; 181:572–579.

8. Bay L, Barnes CJ, Fritz BG, Thorsen J, Restrup MEM, Rasmussen L et al. Universal Dermal Microbiome in Human Skin. mBio 2020; Feb 11:e02945–19.

9. Ganju P, Nagpal S, Mohammed MH, Nishal Kumar P, Pandey R, Natarajan VT et al.Microbial community profiling shows dysbiosis in the lesional skin of Vitiligo subjects. Sci Rep. 2016; 6:18761.

10. Dellacecca ER, Cosgrove C, Mukhatayev Z, Akhtar S, Engelhard VH, Rademaker AW et al. Antibiotics Drive Microbial Imbalance and Vitiligo Development in Mice. J Invest Dermatol. 2020 Mar;140(3): 676-687.e6. doi:10.1016/j.jid.2019.08.435.

11. Dagnelie MA, Corvec S, Saint-Jean M, Nguyen JM, Khammari A, Dreno B. Cutibacterium canes phylotypes diversity loss: a trigger for skin inflammatory process. J Eur Acad Dermatol Venereol. 2019 Dec;33(12):2340–2348. doi:10.1111/jdv.15795. Epub 2019 Aug 20.

12. Rizzatti G, Lopetuso LR, Gibiino G, Binda C, Gasbarrini A. Proteobacteria: A Common Factor in Human Diseases. Biomed Res Int. 2017;2017:9351507. doi:10.1155/2017/9351507. Epub 2017 Nov 2.

13. Li S, Zhu G, Yang Y, Jian Z, Guo S, Dai W, et al. Oxidative stress drives CD8(+) T-cell skin trafficking in patients with vitiligo through CXCL16 upregulation by activating the unfolded protein response in keratinocytes. J Allergy Clin Immunol. 2017 Jul;140(1):177-189.e9. doi:10.1016/j.jaci.2016.10.013. Epub 2016 Nov 5.

14. Nakatsuji T, Chiang HI, Jiang SB, Nagarajan H, Zengler K, Gallo RL. The microbiome extends to subepidermal compartments of normal skin.Nature Comm. 2013; 4:1431.

15. Jakobsson HE, Abrahamsson TR, Jenmalm MC, Harris K, Quince C, Jernberg C et al. Decreased gut microbiota diversity, delayed Bacteroidetes colonisation and reduced Th1 responses in infants delivered by caesarean section. Gut. 2014; 63:559–566.

16. Fadlallah J, Sterlin D, Fieschi C, Parizot C, Dorgham K, Kafsi HE et al. Synergistic convergence of microbiota-specific systemic IgG and secretory IgA. J Allergy Clin Immuno. 2019;l 143:1575–1585.

17. Baurecht H, Ruhlemann MC, Rodrigues E, Thielking F, Harder I, Erkens AS et al. Epidermal lipid composition, barrier integrity, and eczematous inflammation are associated with skin microbiome configuration. J Allergy Clin Immunol. 2018; 141:1668–1676.

18. Opazo MC, Ortega-Rocha EM, Coronado-Arrazola I, Bonifaz LC, Boudin H, Neunlist M et al. Intestinal microbiota influences non-intestinal related auto-immune diseases. Front Microbiol. 2018 Mar 12;9:432. doi:10.3389/fmicb.2018.00432. eCollection 2018.

19. Teulings HE, Limpens J, Jansen SN, Zwinderman AH, Reitsma JB, Spuls PI et al.Vitiligo-like depigmentation in patients with stage III-IV melanoma receivingimmunotherapy and its association with survival: a systematic review and meta-analysis. J Clin Oncol. 2015; 33:773–781.

20. Gopalakrishnan V, Spencer CN, Nezi L, Reuben A, Andrews MC, Karpinets TV, et al.Gut microbiome modulates response to anti-PD-1 immunotherapy in melanoma patients. Science. 2018; 359:97–103.

21. Leonel C, Sena IFG, Silva WN, Prazeres P, Fernandes GR, Mancha Agresti P et al.Staphylococcus epidermidis role in the skin microenvironment. J Cell Mol Med. 2019; 23:5949–5955.

22. Gonzalez T, Biagini Myers JM, Herr AB, Khurana Hershey GK. Staphylococcal Biofilms in Atopic Dermatitis. Curr Allergy Asthma Rep. 2017; 17(12):81.

23. Saint-Georges-Chaumet Y, Edeas M. Microbiota-mitochondria inter-talk: consequence for microbiota-host interaction. Pathog Dis. 2016; 74:ftv096.

24. Yardeni T, Tanes CE, Bittinger K, Mattei LM, Schaefer PM, Singh LN et al. Host mitochondria influence gut microbiome diversity: A role for ROS. Sci Signal. 2019; 12(588):eaaw3159. doi:10.1126/scisignal.aaw3159.

25. Sahoo A, Lee B, Boniface K, Seneschal J, Sahoo SK, Seki T et al. MicroRNA-211 Regulates Oxidative Phosphorylation and Energy Metabolism in Human Vitiligo. J Invest Dermatol. 2017; 137:1965–1974.

26. Dell’Anna ML, Ottaviani M, Kovacs D, Mirabilii S, Brown DA, Cota C et al. Energetic mitochondrial failing in vitiligo and possible rescue by cardiolipin. Sci Rep. 2017; 7:13663.

27. Iacob S, Iacob DG. Infectious Threats, the Intestinal Barrier, and Its Trojan Horse: Dysbiosis. Front Microbiol. 2019; 10:1676.

28. Gilbert JA, Jansson JK, Knight Rl. The earth microbiome project: successes and aspirations. BMC Biol. 2014 Aug 22;12:69. doi:10.1186/s12915-014-0069-1.

29. Bolyen E, Rideout JR, Dillon MR, Bokulich NA, Abnet CC, Al-Ghalith GA et al.Reproducible, interactive, scalable and extensible microbiome data science using QIIME 2. Nat Biotechnol. 2019; 37:852–857.

30. Renaud G, Stenzel U, Maricic T, Wiebe V, Kelso J. deML: robust demultiplexing of Illumina sequences using a likelihood-based approach. Bioinformatics. 2015; 31:770–772.

31. Callahan BJ, McMurdie PJ, Rosen MJ, Han AW, Johnson AJ, Holmes SP. DADA2: High-resolution sample inference from Illumina amplicon data. Nat Methods. 2016; 13:581–583.

32. Katoh K, Misawa K, Kuma K, Miyata T. MAFFT: a novel method for rapid multiple sequence alignment based on fast Fourier transform. Nucleic Acids Res. 2002; 30:3059–3066.

33. Price MN, Dehal PS, Arkin AP. FastTree 2--approximately maximum-likelihood trees for large alignments. PloS One. 2010; 5:e9490.

34. Faith DP. The role of the phylogenetic diversity measure, PD, in bio-informatics: getting the definition right. Evol Bioinform Online. 2007; 2:277–283.

35. Lozupone C, Knight R. UniFrac: a new phylogenetic method for comparing microbial communities. Appl Environ Microbiol. 2005; 71:8228–8235.

36. Bokulich NA, Kaehler BD, Rideout JR, Dillon M, Bolyen E, Knight R et al. Optimizing taxonomic classification of marker-gene amplicon sequences with QIIME 2’s q2-feature-classifier plugin. Microbiome. 2018; 6:90.

37. Quast C, Pruesse E, Yilmaz P, Gerken J, Schweer T, Yarza P et al. The SILVA ribosomal RNA gene database project: improved data processing and web-based tools. Nucleic Acids Res. 2013; 41:D590–596.

38. Mandal S, Van Treuren W, White RA, Eggesbo M, Knight R, Peddada SD. Analysis of composition of microbiomes: a novel method for studying microbial composition. Microb Ecol Health Dis. 2015; 26:27663.

